# Has the COVID-19 pandemic ended or not? opinions from the public in the U.S

**DOI:** 10.1101/2023.02.15.23285880

**Authors:** Yong Yang

## Abstract

Recently President Joe Biden announced the end to the COVID-19 pandemic in the US but some scientists expressed different opinions. This study aimed to examine the view of the end of the COVID-19 pandemic among the public. Data were collected in September 2022 from an online crowdsourcing platform, and respondents answered if they believed that the pandemic has ended in the United States or not. Logistic regressions were used to estimate the likelihood of agreeing on the end of the pandemic, adjusted by demographics and several related variables. Among 2983 respondents, 78.1% believed that the COVID-19 pandemic had ended, and the percentage decreased to 66.5% after adding weights. Males, younger adults, Hispanics, those with higher levels of educational attainment, those with middle levels of household income, those living in suburban or rural areas, and those living in states that voted for the Republican party in the 2020 Presidential Election were more likely to believe that the pandemic had ended, compared with their counterparts. With about one-third of Americans did not agree that the pandemic had ended and marked demographical and geographical differences, the timing and the way of the pandemic end announcement should be deliberately cautious.

## Introduction

After longer than two and half years [1], with more than 6.5 million lives being claimed, the ongoing COVID-19 pandemic may “end in sight” [2]. On September 18, 2022, President Joe Biden announced an end to the COVID-19 pandemic in the US [3]. However, scientists expressed different opinions [4], and some believed that the President’s announcement was premature and harmful to the country’s effort to control COVID-19 [5].

Pandemic, epidemic, and endemic are distinct in theory, however, there are no quantifiable and standard definitions of these terms [4]. The transition from pandemic to epidemic and endemic is a continuous process and the end of a pandemic is subjective to people’s judgment. We argue that most people would agree that a pandemic ends when the disease is under control and cannot disrupt our lives. Here, influenza could be a comparable case. When the risk to health and disruption to daily life from COVID-19 are perceived to be at a similar level to influenza, we may call an end to the COVID-19 pandemic.

Whether the pandemic has ended or not is important because many health, social, and economic policies and regulations could be influenced by the end announcement to some degree. One example is the ongoing COVID-19 public health emergency declaration that has implications for not only COVID-19-related treatments but also many other healthcare services. At the individual level, the end announcement could influence health behaviors such as social distancing, mask-wearing, vaccine uptake, etc.

This study aims to examine the view on the end of the COVID-19 pandemic among the public and the associated factors.

## Methods

Respondents were recruited from Amazon Mechanical Turk (MTurk) [6], an online crowdsourcing platform. Data were collected at the end of September 2022. The Institutional Review Board at the University of Memphis approved this study. For the end of the COVID-19 pandemic, we asked “in your opinion, has the COVID-19 pandemic ended in the United States? by ‘end’, we mean although there are still new infections emerging, these cases are largely under the control, and the spread of coronavirus looks similar to seasonal influenza”. Participants chose either yes or no.

Besides demographics, we collected participants’ residential zip codes, then extracted their residential state, and urbanicity (urban, suburban, rural) [7]. Participants also reported if he/she had ever been infected by COVID-19, physical activity for leisure and for housework using the International Physical Activity Questionnaire (IPAQ) [8], and mental health outcomes including anxiety, depression, and loneliness using the Generalized Anxiety Disorder-2 items (GAD-2) and GAD-7 [9], the Patient Health Questionnaire-2 items (PHQ-2) and PHQ-8 [9], and R-UCLA Loneliness Scale [10].

We estimated the percentage of people who agreed on the end of the pandemic by adding weights to ensure the demographics of the weighted sample match with the US general population by the 2020 Census. A two-step logistic regression modeling was used to estimate the likelihood of agreeing on the end of the pandemic, with the base model including demographical variables, and the final model in which variables including COVID-19 infection, physical activity, and mental health were added to the base model.

## Results

The sample included 2983 participants, and females, older adults, Blacks, those with low educational attainment, and those living in suburban and rural areas were underrepresented in the sample (see Table 1). There were 74.1% of the participants had been infected by COVID-19 during the pandemic, and the majority of participants had mental health disorders, and had a moderate or high level of physical activity. Overall, 2327 participants (78.1%) believed that the COVID-19 pandemic had ended in the US, and the percentage decreased to 66.5% after adding the weights.

**Table 1.**
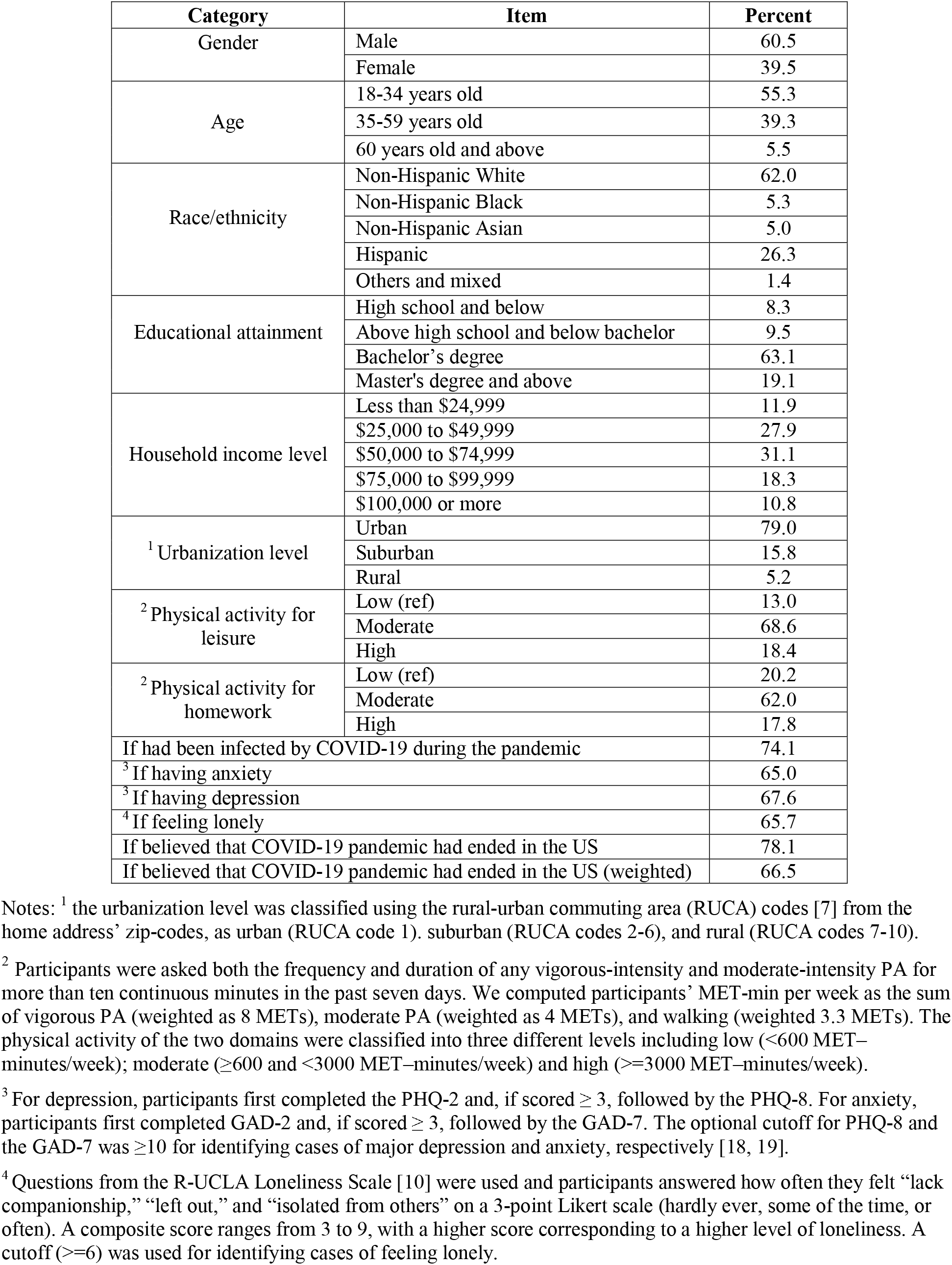
Characteristics of participants (N=2983)

As the results of the baseline model (see Table 2) shows, males, younger adults, Hispanics, those with higher levels of educational attainment, those with middle levels of household income, those living in suburban or rural areas, and those living in states that voted for the Republican party in 2020 presidential election were more likely to believe that the pandemic had ended, compared with their counterparts. In the final model, variables including age groups and educational attainments were not significantly associated with the outcome as they were in the baseline model anymore. Participants who had been infected by COVID-19 during the pandemic (OR=2.00, 95% CI=1.60-2.50), those who had depression (OR=2.38, 95% CI=1.57-3.23), and those who had a moderate (OR=1.90, 95% CI=1.42-2.54) or high (OR=2.37, 95% CI=1.53-3.66) levels of physical activity for leisure were more likely to believe that the pandemic has ended, compared with their counterparts. Several parameters for model fitting indicated that the final model fit data better by adding COVID-19 infection, physical activity, and mental health disorders than the baseline model.

**Table 2.**
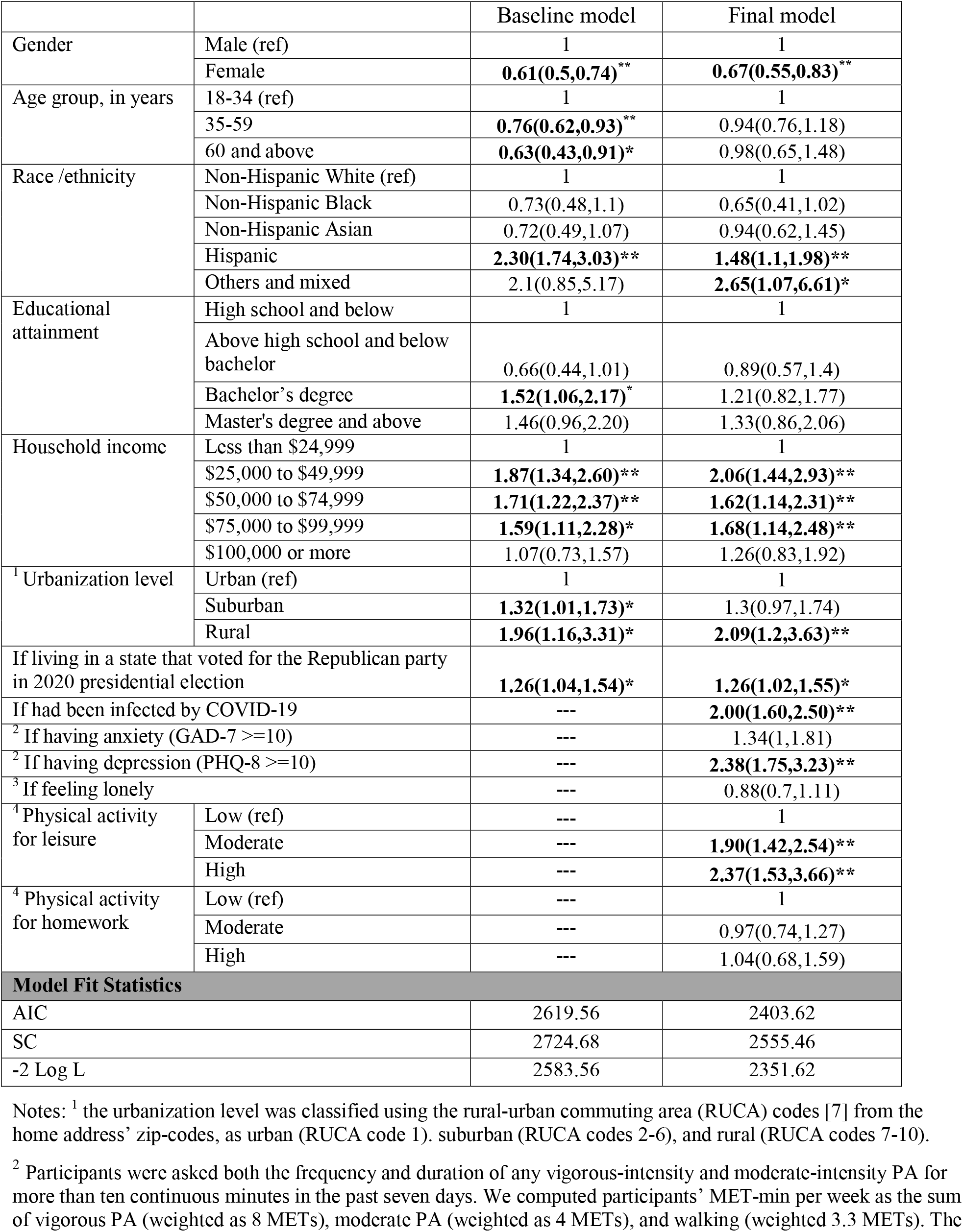
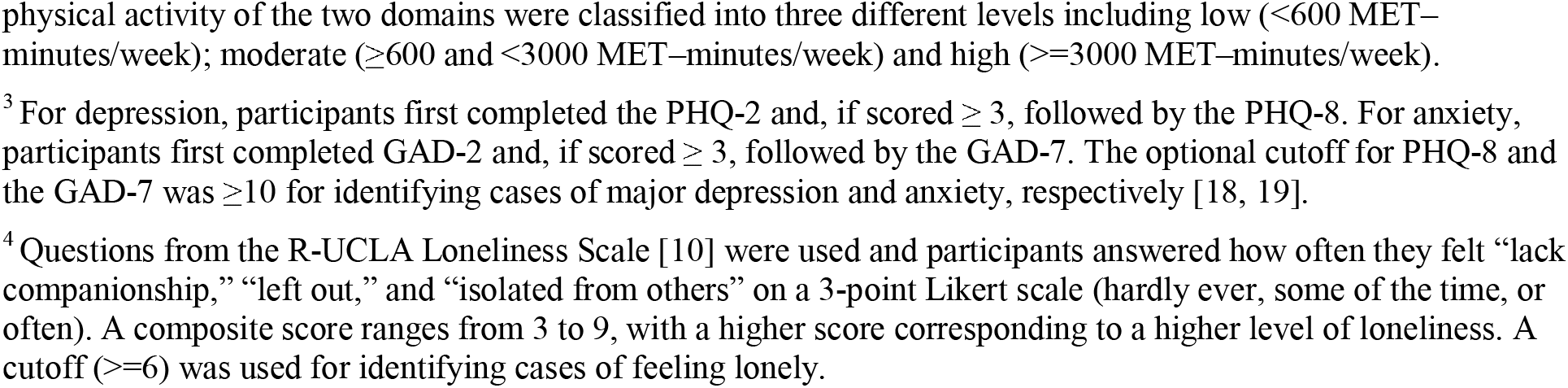
Results (odd ratios) of logistics regression models for the associations between variables and if a participant agreed on the end of the pandemic.

|  |  | Baseline model | Final model |
| --- | --- | --- | --- |
| Gender | Male (ref) | 1 | 1 |
|  | Female | **0.61(0.5,0.74)\*\*** | **0.67(0.55,0.83)\*\*** |
| Age group, in years | 18-34 (ref) | 1 | 1 |
|  | 35-59 | **0.76(0.62,0.93)\*\*** | 0.94(0.76,1.18) |
|  | 60 and above | **0.63(0.43,0.91)\*** | 0.98(0.65,1.48) |
| Race /ethnicity | Non-Hispanic White (ref) | 1 | 1 |
|  | Non-Hispanic Black | 0.73(0.48,1.1) | 0.65(0.41,1.02) |
|  | Non-Hispanic Asian | 0.72(0.49,1.07) | 0.94(0.62,1.45) |
|  | Hispanic | **2.30(1.74,3.03)\*\*** | **1.48(1.1,1.98)\*\*** |
|  | Others and mixed | 2.1(0.85,5.17) | **2.65(1.07,6.61)\*** |
| Educational attainment | High school and below | 1 | 1 |
|  | Above high school and below bachelor | 0.66(0.44,1.01) | 0.89(0.57,1.4) |
|  | Bachelor's degree | **1.52(1.06,2.17)\*** | 1.21(0.82,1.77) |
|  | Master's degree and above | 1.46(0.96,2.20) | 1.33(0.86,2.06) |
| Household income | Less than \$24,999 | 1 | 1 |
| | \$25,000 to \$49,999 | **1.87(1.34,2.60)\*\*** | **2.06(1.44,2.93)\*\*** |
| | \$50,000 to \$74,999 | **1.71(1.22,2.37)\*\*** | **1.62(1.14,2.31)\*\*** |
| | \$75,000 to \$99,999 | **1.59(1.11,2.28)\*** | **1.68(1.14,2.48)\*\*** |
| | \$100,000 or more | 1.07(0.73,1.57) | 1.26(0.83,1.92) |
| <sup>1</sup> Urbanization level | Urban (ref) | 1 | 1 |
|  | Suburban | **1.32(1.01,1.73)\*** | 1.3(0.97,1.74) |
|  | Rural | **1.96(1.16,3.31)\*** | **2.09(1.2,3.63)\*\*** |
| If living in a state that voted for the Republican party in 2020 presidential election |  | **1.26(1.04,1.54)\*** | **1.26(1.02,1.55)\*** |
| If had been infected by COVID-19 |  | --- | **2.00(1.60,2.50)\*\*** |
| <sup>2</sup> If having anxiety (GAD-7 >=10) |  | --- | 1.34(1,1.81) |
| <sup>2</sup> If having depression (PHQ-8 >=10) |  | --- | **2.38(1.75,3.23)\*\*** |
| <sup>3</sup> If feeling lonely |  | --- | 0.88(0.7,1.11) |
| <sup>4</sup> Physical activity for leisure | Low (ref) | --- | 1 |
|  | Moderate | --- | **1.90(1.42,2.54)\*\*** |
|  | High | --- | **2.37(1.53,3.66)\*\*** |
| <sup>4</sup> Physical activity for homework | Low (ref) | --- | 1 |
|  | Moderate | --- | 0.97(0.74,1.27) |
|  | High | --- | 1.04(0.68,1.59) |
| **Model Fit Statistics** |  |  |  |
| AIC |  | 2619.56 | 2403.62 |
| SC |  | 2724.68 | 2555.46 |
| -2 Log L |  | 2583.56 | 2351.62 |

Notes: <sup>1</sup> the urbanization level was classified using the rural-urban commuting area (RUCA) codes [7] from the home address' zip-codes, as urban (RUCA code 1), suburban (RUCA codes 2-6), and rural (RUCA codes 7-10).
<sup>2</sup> Participants were asked both the frequency and duration of any vigorous-intensity and moderate-intensity PA for more than ten continuous minutes in the past seven days. We computed participants' MET-min per week as the sum of vigorous PA (weighted as 8 METs), moderate PA (weighted as 4 METs), and walking (weighted 3.3 METs). The
physical activity of the two domains were classified into three different levels including low (<600 MET–minutes/week); moderate ( $\geq 600$ and <3000 MET–minutes/week) and high ( $\geq 3000$ MET–minutes/week).
<sup>3</sup> For depression, participants first completed the PHQ-2 and, if scored $\geq 3$ , followed by the PHQ-8. For anxiety, participants first completed GAD-2 and, if scored $\geq 3$ , followed by the GAD-7. The optional cutoff for PHQ-8 and the GAD-7 was $\geq 10$ for identifying cases of major depression and anxiety, respectively [18, 19].
<sup>4</sup> Questions from the R-UCLA Loneliness Scale [10] were used and participants answered how often they felt “lack companionship,” “left out,” and “isolated from others” on a 3-point Likert scale (hardly ever, some of the time, or often). A composite score ranges from 3 to 9, with a higher score corresponding to a higher level of loneliness. A cutoff ( $\geq 6$ ) was used for identifying cases of feeling lonely.

## Discussion

Overall, our results indicate that the majority (about two third) of the US population believed that the COVID-19 pandemic had ended. This is understandable considering the much lower numbers of new cases and deaths over the past several months compared with the several peak times, most Americans have received at least one dose of COVID-19 vaccine, and many restrictions such as social distancing were relaxed in many places, and most people do not wear masks anymore. However, the statistics of COVID-19 are still dire, one major reason that the announcement of the pandemic end was perceived as “premature”. During the last week of September 2022, on average, each day saw about 50000 new cases and above 400 deaths due to COVID-19. This inconsistency may reflect that with the pandemic running much longer than most people expected and people may be habituated to the situation, and people’s desire to back to a “normal life” is strong enough and people would rather tolerate a higher level of risk than what people generally accept before the pandemic.

The different opinions for the end of the pandemic may be explained by at least three reasons. First, people whose health and life had been impacted more during the pandemic may be more likely to agree with the end of the pandemic after the negative impact was relieved, for example, those who had been infected and then recovered from the COVID-19 compared with those who had never been infected. Participants with middle levels of household income were more likely to agree with the end of the pandemic and a possible reason is that although both middle and low levels income groups were more likely to endure financial hardship at the beginning of the pandemic [11], people with middle income have higher resilience to recover over time compared with those with low levels of income. The second reason is politics. Those living in states that voted for the Republican party in the 2020 presidential election and those living in suburban or rural areas were more likely to be Republicans or lean towards the republication party, and they were more likely to believe the pandemic was exaggerated [12, 13]. Third, some groups tend to be more optimistic than others. For example, we found males and Hispanics were more likely to believe at the end of the pandemic. This is consistent with some evidence that males and Hispanics are more optimistic than their counterparts for various issues [14-16], and importantly, a recent study conducted at the end of 2021, reported that males were more optimistic about the ending of the pandemic than females and Hispanics were more optimistic for the ending of the pandemic than other racial/ethnical groups [17]. Interestingly, we found physical activity and depression were significantly associated with the outcome. Probably people who believe that the pandemic is still ongoing tend to limit their physical activity for leisure at least outdoors. Unfortunately, we could not provide a plausible explanation of why depression but not anxiety and loneliness were associated with people’s opinions at the end of the pandemic, and further studies may be needed.

Our findings may provide implications for COVID-19 policies. With about one-third of Americans did not agree that the pandemic has ended and marked demographical and geographical differences, the timing and the way of the pandemic end announcement should be deliberately cautious. For example, it may take several steps at the federal level, follow and combine with various levels of epidemic and endemic at state and local levels because COVID-19 is unlikely to disappear in a short term, but rather, will keep emerging somewhere occasionally, sometimes serious, similar to influenza. Indeed, whether the pandemic has ended or not, the mitigation effort to reduce the negative influence of the pandemic will continue for a long period.

One limitation of this study is that our sample is not representative of the US population, although weights were added to match the US general population for major demographics, some groups, for example, people who have no access to the Internet or are illiterate were excluded from the survey. Another limitation is that our study did not collect information about how various aspects of daily life was impacted by the COVID-19 pandemic that may help to explain people’s different view on the end of the pandemic.

## Data Availability

All data produced in the present study are available upon reasonable request to the authors

## Notes

### Competing Interest Statement

The authors have declared no competing interest.

### Author Declarations

The Institutional Review Board at the University of Memphis approved this study.

